# Quantitative and geometric motor unit analysis using magnetomyography

**DOI:** 10.1101/2023.06.09.23291204

**Authors:** Philip J. Broser, Thomas Middelmann, Nima Noury, Markus Siegel, Stefan Hartwig, Thomas Klotz, Justus Marquetand

## Abstract

**Objective:** Magnetomyography (MMG) is currently a rather unexplored neurophysiological modality and it is not known to which extent the number of motor units have an influence on the amplitude and the direction of the MMG-signal.

**Methods:** A simultaneous invasive electromyography (iEMG), surface EMG (sEMG) and MMG using optically pumped magnetometer (OPM-MMG) of the right abductor digiti minimi muscle (ADM) of two healthy participants was recorded during a stepwise increasing electrical stimulation of the ADM innervating ulnar nerve. Then, the number of electrically evoked motor units was estimated (MUNE), the magnetic field vectors were reconstructed and aligned to the muscular anatomy. In addition, a finite element simulation of the ADM muscle was performed and compared to the experimental data.

**Results:** The more motor units were activated by increasing electrical stimulation, the stronger the MMG signal became, which was the same for iEMG&sEMG (r>0.96). The finite element simulation showed the same relation between the magnetic and electric signal. Further, based on the simulation the number of activated muscular fibers and neuromuscular units could be estimated the ratio of signal to fibers determined.

In addition, the precise vector direction of the magnetomyography (MMG) signal can reliably be recorded following the electric stimulation of the ulnar nerve and followed the muscle fiber direction.

**Conclusion:** The MMG signal can be used to determine the amount of activated motor units, but also analysis of the magnetic field vector corresponds to the muscle fiber direction, offering a functional as well as structural characterization of muscles. The modelling and simulation is especially helpful to understand the magnetic muscular signal in detail.

**Significance:** Next to establishing MUNE in MMG, our results provide the first quantitative comparison between MMG vs. iEMG&sEMG and highlight the possibilities of the vector component analysis in MMG.

**Highlights:**

- Comparative study of MMG, iEMG&sEMG using electrically induced activation of motor units.
- MUNE in MMG is possible and is potentially superior to surface EMG.
- The vector components of the MMG-signal correspond to the muscle fiber direction of the muscle.
- Finite element simulation of the muscular magnetic and electric signal

## 1 Introduction

The control of muscle force is regulated by the firing rate of the motor neurons and the number of motor units activated. The number of motor units activated vary depending on the motor task an individuum performs (Heckmann, 2012) and decrease in old age as well as in motor neuron diseases such as amytrophic lateral sclerosis (ALS) or spinal muscular atrophy (SMA). Therefore, the number of motor units represents a biomarker for the integrity of the neuromuscular system as well as for any progression in motor neuron diseases. Although the exact measurement of the number of motor units in a muscle in vivo is impossible, several electrophysiological techniques exist to provide an estimate; one, conventionally used the daily clinical routine is the so-called motor unit number estimation (MUNE). Here, a peripheral nerve innervating a muscle will be electrically stimulated with small, stepwise increments, which leads to small and stepwise increases of the compound muscle action potential (CMAP), which then can be measured in the recording area of surface electromyography (EMG)-electrodes. In principle, in MUNE, the CMAP-amplitude, -duration or -area will be then divided by the average amplitude, duration or area of a single motor unit action potential (SMUP). Despite different MUNE techniques and principal inaccuracy, MUNE has been proven repeatedly in clinical studies to be capable of monitoring motor neuron loss in various neuromuscular diseases with surface EMG.

Considering the magnetic pendant of EMG, magnetomyography (MMG), with all its ad advantages and disadvantages (for a comprehensive overview, we refer to our previous work), it remains an open question if and how similar MMG can also be utilized for MUNE. To date, MMG has scarcely been explored regarding a clinical-neurophysiological application, making it reasonable to further investigate MMG in this field. Specifically, the possibility to measure the magnetic muscle signal contactless using miniaturized quantum sensors (so-called optically pumped magnetometers, OPM) seems to be a potential practical advantage over surface EMG. Furthermore, the ability to quickly and easily determine the magnetic field vector also appears to be an interesting feature of MMG as the field vector could not only be used for the measurement of signal strength, but also for the estimation of the muscle fiber direction.

Consequently, we investigated MUNE in MMG of the abductor digiti minimi muscle (ADM) and compared it with simultaneously recorded invasive and surface EMG. In order to understand the signal generation in more detail we performed a finite element simulation of the propagating muscular action potential. MMG offers the unique opportunity to measure not only the signal strength but also the vectorial direction of the magnetic signal. To investigate the potential of this information to support MUNE, a lidar scan of the experimental setup was performed and the magnetic field vectors were put in relation to the precise anatomical structures.

## 2 METHOD

### 2.1 Principal setup

A two subject, experimental study was conducted at the PTB in the BSMR2 magnetic shielded room at the PTB in Berlin, Germany. The data from both recorded subjects could be used for further analysis. The data of subject one is presented in great detail in the result section. The data of subject two is included as supplementary data. Experiments were performed-according to the standards set by the World Medical Association (World Medical Association, 2001). The subjects gave their consent for the data to be published and the subjects are authors of this study. The experiment was designed specifically to record the magnetic activity of the right abductor digiti minimi muscle (ADM, Figure 1) after electric stimulation of the supplying nerve (Figure 1 Panel A). In order to stimulate the axons of the motor neurons of the ulnar nerve, the nerve was stimulated with two superficial electrodes at the ulnar fossa. For experiment preparation the subjects sat down on a comfortable chair inside the magnetic shielded room (BSMR2, PTB, Berlin). The right hand and arm of the subject was supported by pillows (Figure 1 B) so that the hand rested completely relaxed on the table. Two standard non magnetic ECG electrodes (as previously used, compare with Broser, 2018) were placed on the ulnar fossa at the level of the elbow after cleaning the skin with alcohol and a soft rub to reduce the skin conductive resistance. The stimulation cables were tunneled outside the magnetic shieled room with two 5m long nonmagnetic wires and connected to the electric stimulator (Micromed, Venice, Italy) outside the shieled room. In addition to the stimulation impulse, the stimulator sends a trigger signal to the recording apparatus in order to synchronize the recording. The recording was performed with a monopolar pulse of 0.1ms duration and a systematic varying current strength from (7.5 mA to 20 mA).

**Figure 1.**
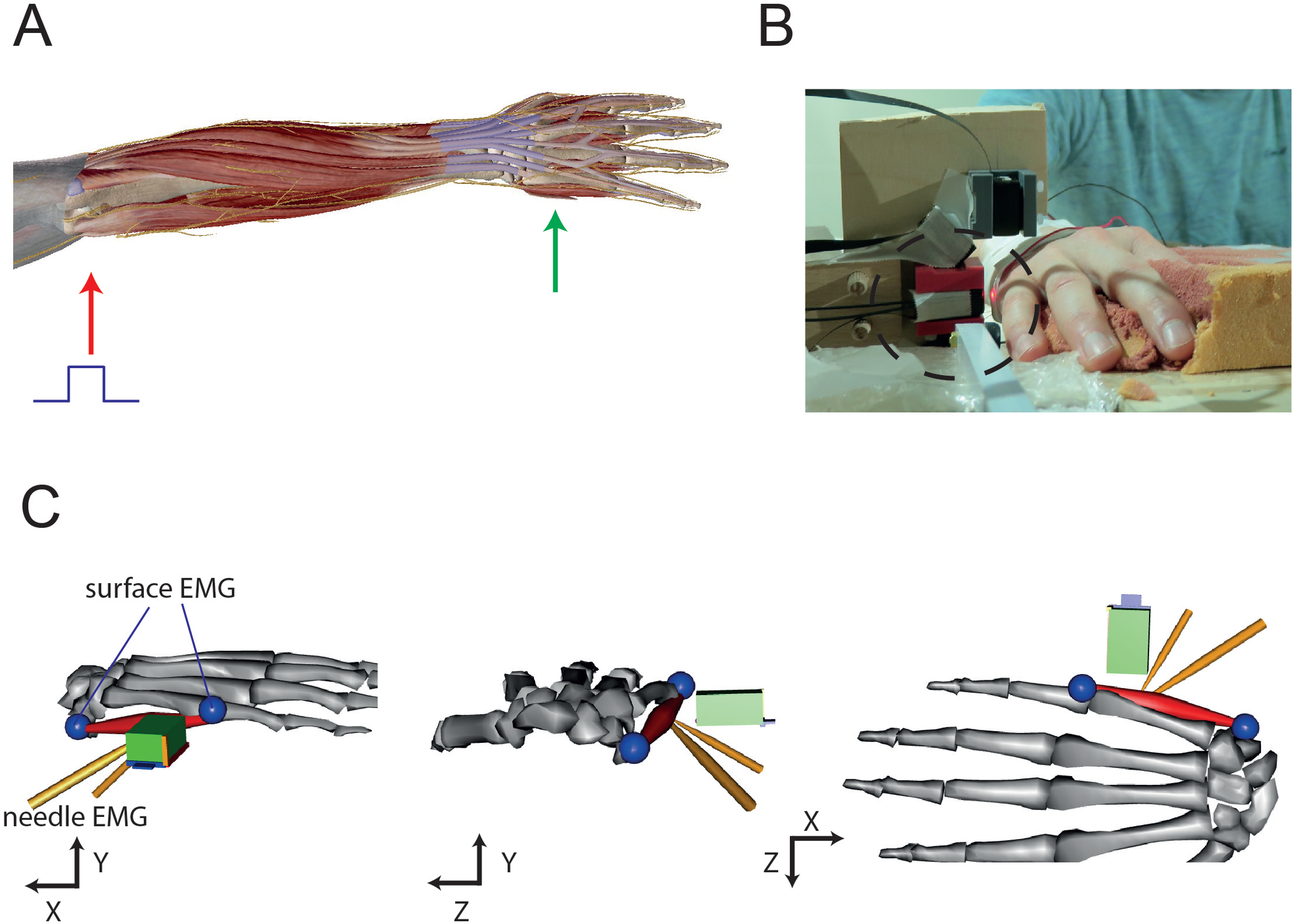
Panel A: Anatomical image showing the stimulation location at the ulnar fossa (red arrow) and the recording location at the site of the musculus abductor minimi (green arrow) Panel B: Photograph of the subject and the location of the relevant OPM sensor (black dotted circle) Panel C: Hand modell with the location of the - surface EMG electrodes – blue spheres - fine wire needle EMG electrodes – yellow cylinder - OPM sensor – green cuboid (blue – x-direction, yellow – y-direction, red – z-direction)

The electric muscle activity was recorded by two monopolar^1^ fine-wire EMG needles (MedCat 0,4 x 10 mm Platin-Iridium needle) which were placed inside or near the ADM and two surface electrodes (Figure 1 C) which were placed at the palmar side of the thenar region.

Five optically pumped magnetometers (Generation QZFM-gen-3, sensitive for all three spatial directions x,y,z, QuSpin Inc., Louisville, CO, USA) were placed in a circular order around the muscle (see Figure 1 B). However, for the study presented here only the main OPM sensor normal to the skin and muscle longitudinal extension (see Figure 1C) was used.

The magnetometers were placed about 5 - 10 mm above the skin surface.

The sensors are based on optically detecting zero-field resonance in hot rubidium vapor, which was contained in a vapor cell measuring 3 × 3 × 3 mm^3^. The small size of the OPM sensors allows for easy handling and their flexible adaptation to specific geometrical situations (Boto et al., 2017, Osborne et al., 2018, Sander et al. 2020). OPMs are capable of measuring three components of the magnetic field vector: the x-, y- and z-direction. They measure with a magnetic field sensitivity in the order of 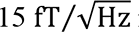 in a bandwidth of 3–135 Hz and a dynamic range of a few nanoteslas. To adapt to a non-zero magnetic background field, the sensors are equipped with internal compensation coils that can cancel magnetic background fields of up to 200 nT in the sensing vapor cell.

### 2.2 Recording

The signal from the OPM devices was recorded with the data acquisition system of the BSMRII (LANDAQ, Berlin) which is permanently installed in the magnetically shielded rom. In addition, the signal from the two monopolar and two surface EMG were recorded with a NeurOne-EEG high quality Neurophysiology recording system. To synchronize both systems the TTL signal from the electric stimulator was fed into both the LANDQ and the NeurOne-EEG recording system. This trigger was used to time register both recorded datasets and to mark the trial onsets.

The analogue signals from the OPMs were digitized with a sampling rate of 8000 samples per second. A band pass filter with a high-pass of 10 Hz and a low-pass filter of 300 Hz was applied to the analogue signal of the OPMs.

The EMG / EEG was recorded with 20 kilo samples per second and filtered with a butterworth filter with a high-pass of 60Hz and a low-pass filter of 950 Hz.

The OPM system has an intrinsic delay between the measurement and analogue output; this delay was measured to be 3.5 ms. In addition the ADC system of both signal types and the filter introduces a further delay of 2.5ms. Based on the trigger artifact visible in the OPM and EMG, the data was post-hoc calibrated so that the magnetic field, electric potential, and signal of the stimulator were in temporal synchrony.

### 2.3 Data processing and simulations

As described above, the data was recorded with the LANDAQ and NeuroOne-EEG System. The LANDAQ data was exported in the proprietary continuous float file format and imported to R (R Core Team, 2022), for further processing the NeuroOne-EEG data was exported to the EDF File Format and also imported into R. Based on the trigger signal both datasets were temporally reregistered and stored as a single data object in R. The calibration formula given by the manufacturer of the OPM sensor (Quspin, Boulder Colorado) was used to convert the recorded ADC values into field strength in Tesla.

### 2.4 Geometric reconstruction – LIDAR scan

To precisely reconstruct the location and orientation of the OPM sensors in relation to the anatomical structures a LIDAR scan (XXXX) was performed at the beginning of the experiment. The lidar scanner generated a 3d mesh object of the precise arrangement of the sensor array and the subject (see Supplementary Figure 1A). The 3d mesh object from the LIDAR scanner was exported as obj File and imported into R using the rgl package (Murdoch, 2021). The anatomical data of the OpenSim Package was used to generate a realistic structural model of the hand of the subject and its relation to the OPM sensors (Figure 1 C and Supplementary Figure 1 B) (Seth et al. 2018).

### 2.5 Finite Element Simulation of the propagating muscular action potential

The finite element simulation was done as described in (Klotz 2022).

## 3 Results

Subject one has an lower arm length of 23 cm (elbow to writs). As expected for a person with a regular nerve conduction speed, about 8ms after electric stimulation with 12mA at the ulnar fossa, a MMG (Figure 2 Panels A to C) and EMG (Figure 2 Panels E to G) signal could be recorded from the abductor digiti minimi muscle. As shown in Figure 1 the OPM sensor and the fine wire EMG needle were placed slightly towards the distal end of the ADM muscle. Therefore both the OPM sensor but also the EMG electrodes pick up more boundary effects to the muscular activity leading to a polyphasic signal (compare with Figure 2 panels C and E).

**Figure 2.**
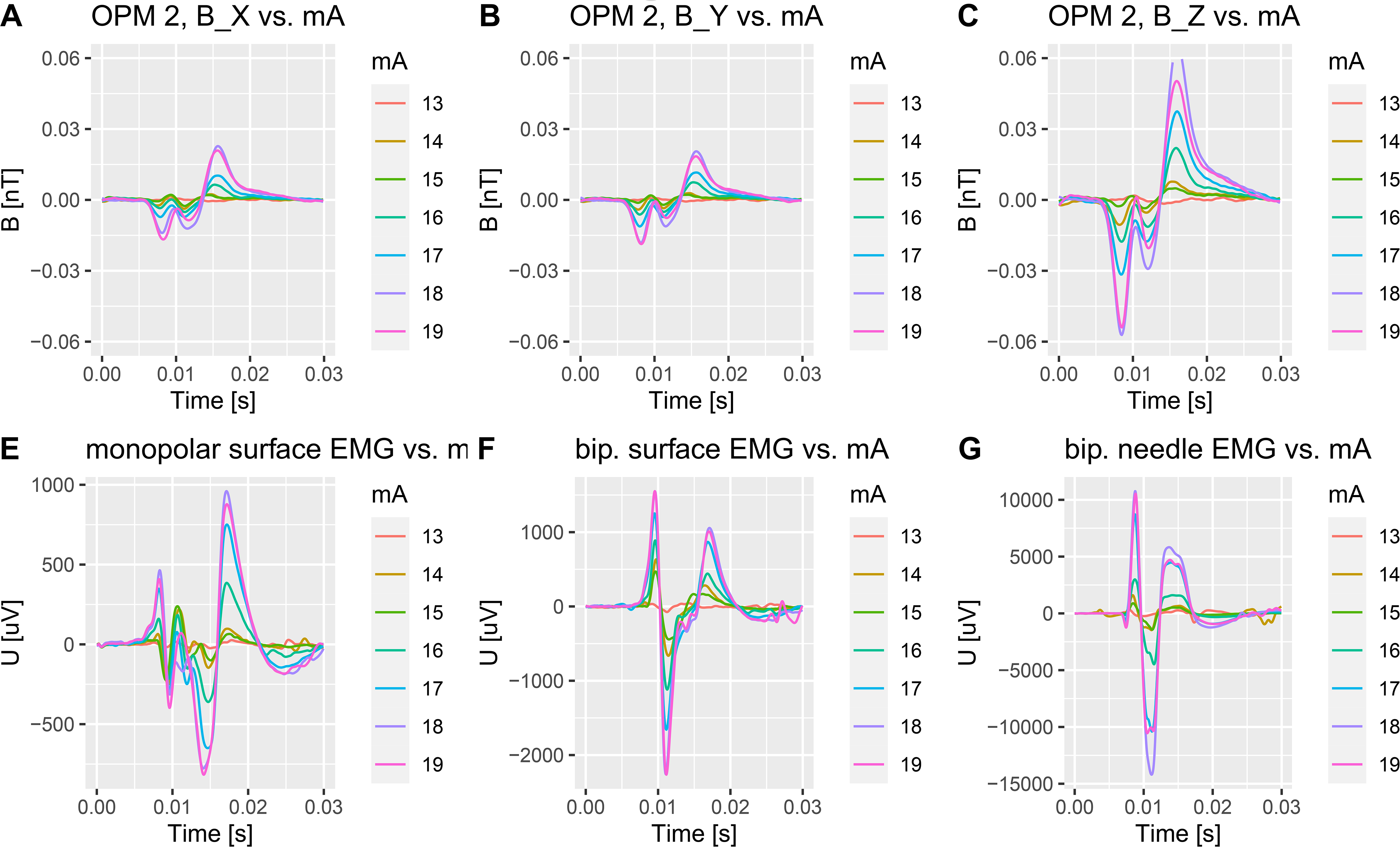
Panels A to C – Signal of the OPM in the different directions. Each trace belongs to a single stimulation strength from 13mA to 19mA (right side) Panel E: Signal of the distal surface electrode versus a common ground electrode Panel F: Signal of the bipolar surface electrode Panel G: Signal of the bipolar fine wire electrodes

### 3.1 MMG and EMG signal magnitude in relation to the number of active neuro muscular units

The ulnar nerve consist of about 40000 axonal fibers with 2670 (+-350) being efferent axons mainly axons of motor neurons (Gesslbauer et. Al 2017). Of these motor neurons about 130 innervate the ADM muscle (Gawel, 2014). Each of these motor neuron axons innervates about 100 ADM muscle fibres and form a neuro muscular unit. Depending on the diameter of these axons they are activated at different stimulation levels. To see, how the MMG signal changes with increasing numbers of activated neuro muscular units the electric stimulation strength was systematically increases from 11 mA (the minimum response intensity) to 20 mA (supra maximal response).

As visible in Figure 2 the signal from the OPM and the recorded EMG signal (needle and surface) increases with increasing stimulation strength until the amplitude reaches an plateau with about 19mA (see Figure 2 and Table 1).

**Table 1.** For each stimulation strength the min-max amplitude for each modality was calculated (left part) and in addition for each modality the area under the curve for each modality was calculated.

| Stim [mA] | Min-Max Amplitude |  |  |  | Area under the curve |  |  |  |
| --- | --- | --- | --- | --- | --- | --- | --- | --- |
|  | OPM2_Z | EMG1 | dsEMG | dnEMG | OPM2_Z_Int | EMG1_Int | dsEMG_Int | dnEMG_Int |
| 11 | 0.00172 | 50.36 | 79.27 | 1,195.66 | 0.04489 | 2,251.33 | 4,523.93 | 54,534.14 |
| 12 | 0.00275 | 67.06 | 87.86 | 611.28 | 0.08490 | 4,275.28 | 5,300.37 | 32,199.25 |
| 13 | 0.00345 | 53.16 | 119.25 | 1,110.18 | 0.09223 | 3,828.09 | 6,297.87 | 58,686.10 |
| 14 | 0.01826 | 471.59 | 1,296.96 | 3,075.33 | 0.59765 | 22,625.50 | 54,905.15 | 155,862.62 |
| 15 | 0.00843 | 461.85 | 913.51 | 2,324.16 | 0.33345 | 20,317.10 | 46,825.85 | 108,983.96 |
| 16 | 0.03965 | 746.51 | 2,007.40 | 7,472.52 | 1.25092 | 53,834.12 | 85,995.74 | 387,708.28 |
| 17 | 0.06906 | 1,400.91 | 2,910.33 | 19,133.62 | 2.26655 | 99,390.40 | 152,618.22 | 985,016.45 |
| 18 | 0.12301 | 1,737.36 | 3,803.02 | 24,963.25 | 3.86482 | 123,423.50 | 186,162.14 | 1,277,650.29 |
| 19 | 0.10429 | 1,694.62 | 3,815.86 | 21,076.96 | 3.06896 | 120,318.62 | 180,390.94 | 1,078,918.10 |
| 20 | 0.07411 | 1,132.76 | 2,831.46 | 12,302.61 | 2.15832 | 84,289.46 | 124,998.54 | 679,650.85 |

For all stimulation strengths the signal magnitude is highest in the sensor Z direction (orthogonal to the direction of the muscle and orthogonal to the skin) which fits with the simulation data from the finite element model by (Klotz, 2022).

### 3.2 Quantitative Analysis of the signal in relation to stimulation strength and in comparison to the EMG

In order to quantify the MMG and EMG signal strength the negative to positive peak amplitude and the area under the curve (absolute integral) was calculated for the different stimulation strength (Table 1). All Signal Types (OPM, surface and needle EMG) show an increase in signal amplitude with increasing stimulation strength. With around 11 mA of stimulation strength a stable all or nothing response can be triggered in all recording modalities. The recorded magnetic signal and the electric signal increase in synchrony. This is especially evident when plotting the magnetic signal in relation to the different electrically recorded signals (see Figure 3). Both the needle and the surface EMG show a linear relation with the magnitude of the magnetic signal in all three spatial directions. The correlation is highly significant for the surface and needle emg with the OPM signal (R>0.9, p<0.01).

**Figure 3.**
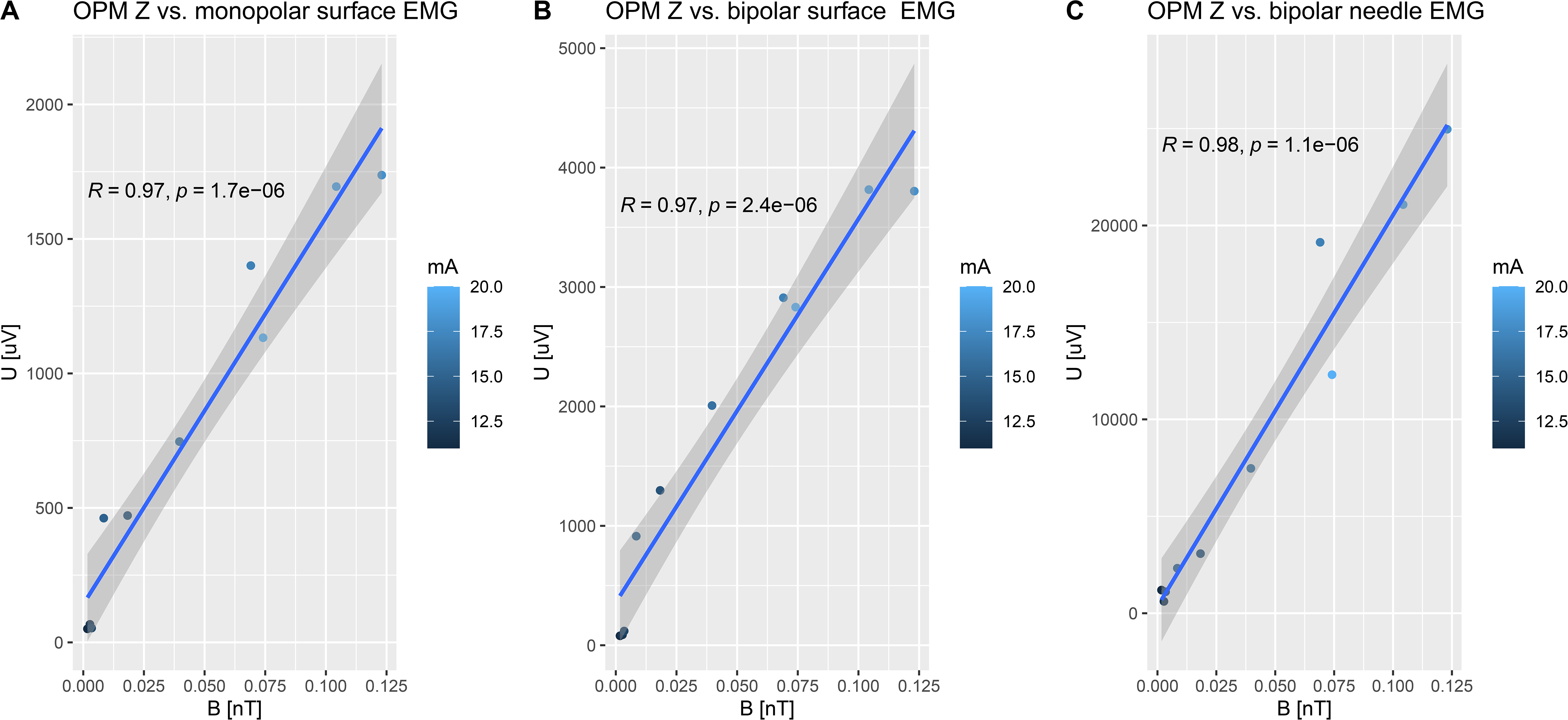
Panel A: Correlation plot of the MMG Signal in Z direction (x-Axis) versus the surface monopolar EMG signal. Panel B: Correlation plot of the MMG Signal in Z direction (x-Axis) versus the surface bipolar EMG signal. Panel C: Correlation plot of the MMG Signal in Z direction (x-Axis) versus the needle bipolar EMG signal.

### 3.3 Estimation of the number of neuro muscular units

In order to estimate the number of neuro muscular units it is necessary to estimate the smallest stimulation with an all or nothing response. Based on the projections shown in Figure 3 this was estimated to be at 11mA. Table 2 shows the amplitude min max and the area under the curve (integral) for the OPM and EMG data. The last row shows the ration between the minimal signal and maximum signal for each modality (ration 11 to 19mA). The ratio of the minimal and maximal signal is given in the last row. A so the estimate of the number of neuro muscular units differs between about 40 and 80.

**Table 2.** The signal from the minimal stimulation with an all or nothing response (11mA) and the supra maximal stimulation response (19mA) was used to estimate the number of motor units.

| Stim [mA] | Min-Max Amplitude |  |  |  | Area under the curve |  |  |  |
| --- | --- | --- | --- | --- | --- | --- | --- | --- |
|  | OPM2_Z | EMG1 | dsEMG | dnEMG | OPM2_Z_Int | EMG1_Int | dsEMG_Int | dnEMG_Int |
| 11.000000 | 0.00172 | 50.36 | 79.27 | 1,195.66 | 0.04489 | 2,251.33 | 4,523.93 | 54,534.14 |
| 19.000000 | 0.10429 | 1,694.62 | 3,815.86 | 21,076.96 | 3.06896 | 120,318.62 | 180,390.94 | 1,078,918.10 |
| 1.727273 | 60.61678 | 33.65 | 48.13 | 17.63 | 68.37177 | 53.44 | 39.87 | 19.78 |

### 3.4 Simulation of the magnetic and electric signal

Figure 4 shows the magnetic field vectors (Panels A to C) and electric signals (Panels E and F) obtained by a finite element simulation of a model of the ADM. The muscular anatomy and the positions were modelled according to the experiment. During the modelling the number of active neuro muscular units and thus muscle fibres were systematically increased. Comparable to the experiment the amplitude increases but the shape of the signal remains unaltered. Interestingly when plotting the area under the curve for the magnetic and electric signal one obtains again a linear relation. Figure 4 Panel G shows the relation of the electric and magnetic signal.

**Figure 4.**
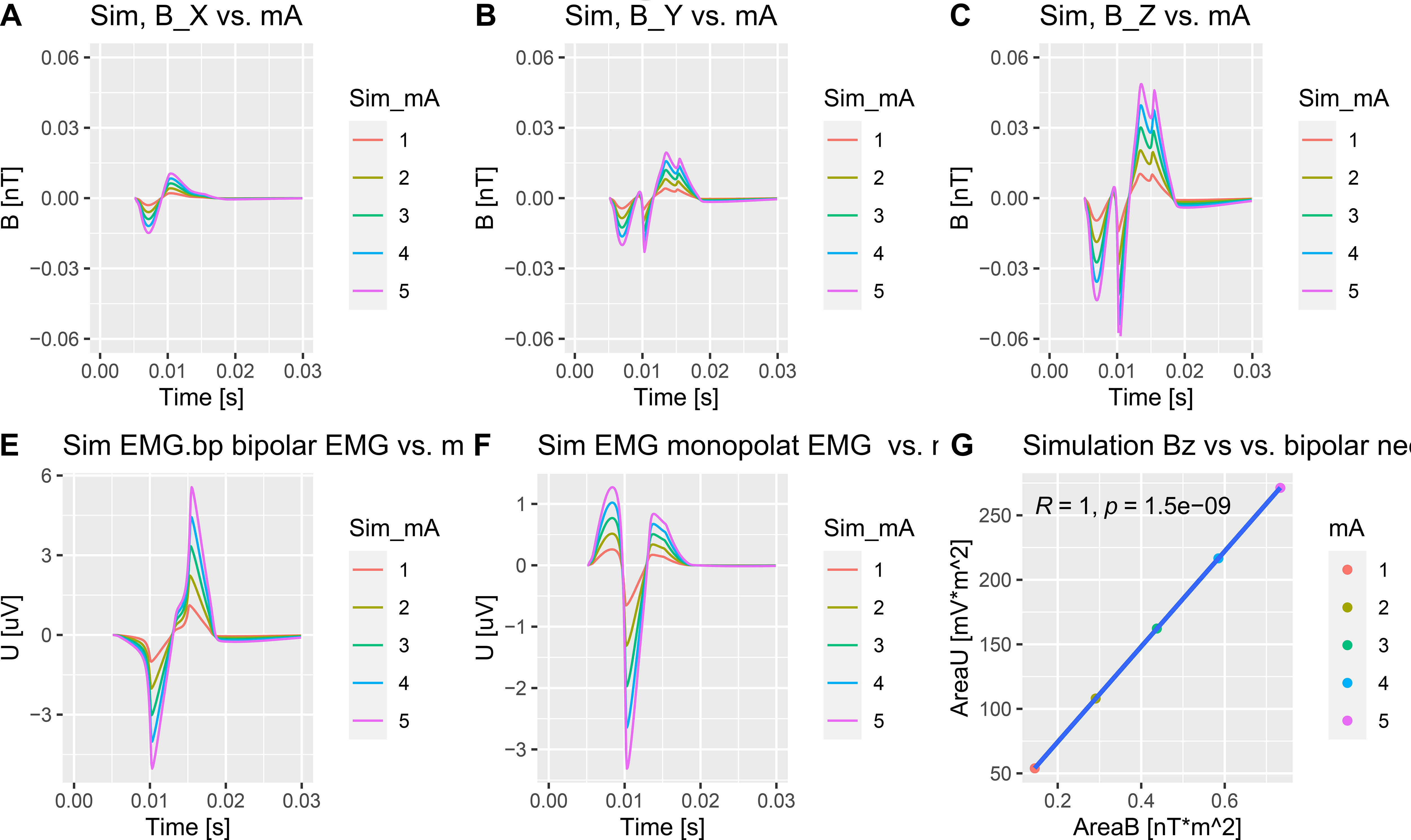
Panel A to C: Simulated Magnetic field vectors Panel E: Simulated bipolar surface EMG Panel F: Simulated donopolar surface EMG Panel G: Correlation of the simulated magnetic and electric signal strength (Color reflects the numbers of neuro muscular units active in the simulation)

### 3.5 Direction of the signal vector

The third generation QSpin OPMs are sensitive in all three spatial directions. Therefore, the magnetic field vector can be reconstructed. Figure 5 Panel A shows the signal trace of the z direction with the location of the negative maxima marked by a blue vertical line and the positive maxima for each trace marked by a red vertical line. The resulting vectors taking all three directions into consideration are shown in Panel B. For better visualization the vectors are moved next to the OPM sensor. The direction of the negative maxima and positive maxima stays almost constant with the increasing stimulation strength. Indeed table 3 shows that the variability of the angle of the minimal vector has a standard deviation of the angle to the X axis of only 5° with Y and Z being even lower. The variability of the direction of the positive vector is even lower. Overall the direction of the magnetic field vectors stay stable over a large range of stimulation intensities.

**Figure 5.**
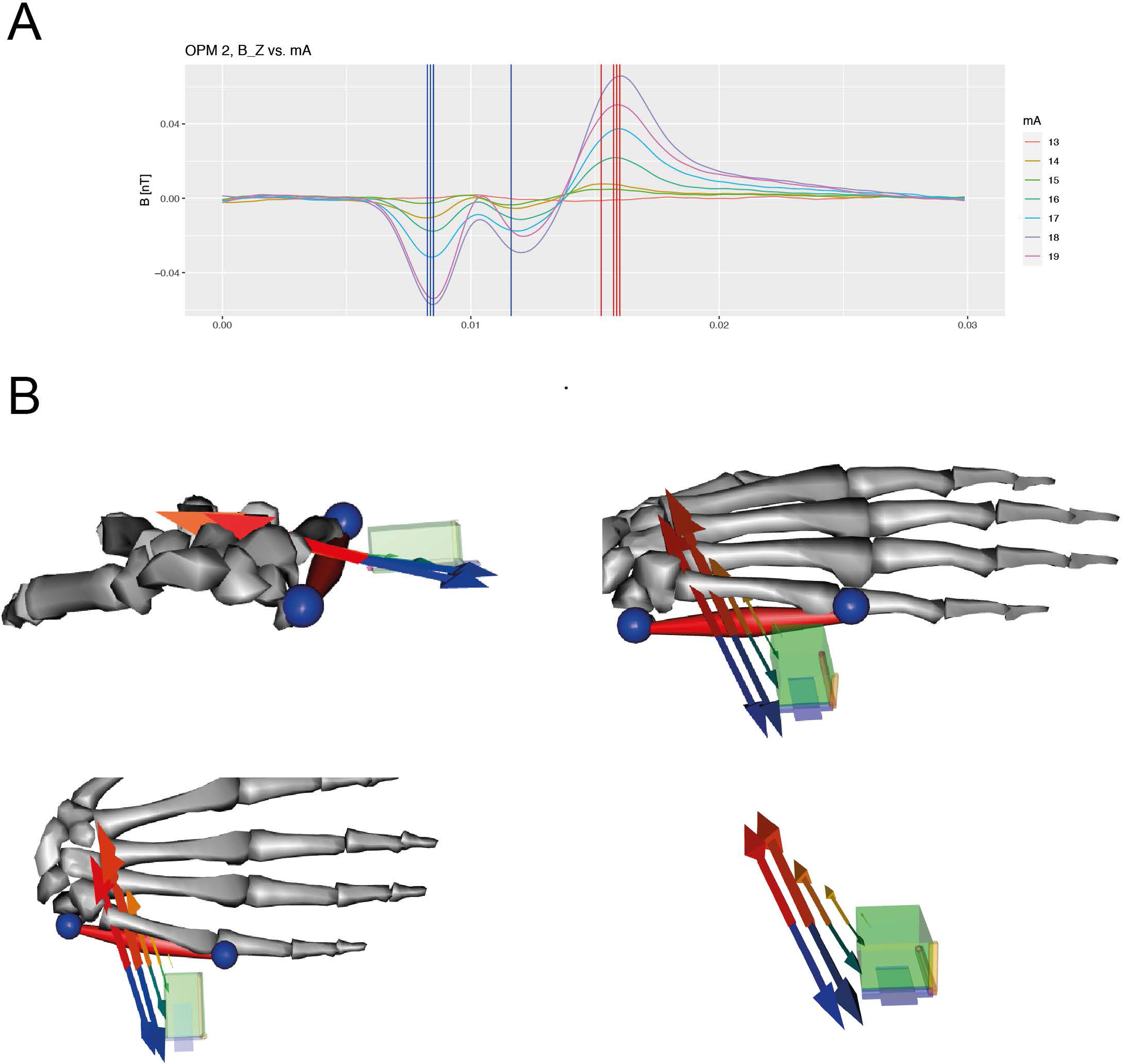
Panel A: MMG signal in Z direction for different stimulation strength plotted with different colors. The local minima for each trace is marked by a blue vertical line and the local maxima is marked by a red vertical line. Panel B: Graphical representations of the vectors of the minimum location (different shades of blue) and maximum locations (different shades of red). The vectors are drawn with a slight offset to the sensor to enhance visibility.

**Table 3.** For each stimulation from 13mA to 19mA the angle to of the minimal and maximal magnetic field vector against the different cartesian axis (X,Y,Z) was calculated. The mean and standard deviation of the directions are given in the last two rows.

|  |  | Min Vector |  |  | Max Vector |  |  |
| --- | --- | --- | --- | --- | --- | --- | --- |
|  | Stim [mA] | X | Y | Z | X | Y | Z |
|  | 13 | 100.2 | 102.9 | 163.5 | 77.1 | 73.0 | 21.6 |
|  | 14 | 94.8 | 106.9 | 162.4 | 76.7 | 73.8 | 21.2 |
|  | 15 | 117.0 | 112.0 | 143.9 | 78.8 | 75.2 | 18.7 |
|  | 16 | 95.7 | 105.9 | 163.0 | 75.7 | 73.0 | 22.5 |
|  | 17 | 100.1 | 107.7 | 159.4 | 75.6 | 73.8 | 22.0 |
|  | 18 | 101.9 | 105.9 | 160.0 | 72.3 | 74.0 | 24.3 |
|  | 19 | 106.1 | 107.5 | 155.9 | 69.0 | 71.6 | 28.5 |
| avg | — | 102.25 | 106.97 | 158.30 | 75.02 | 73.48 | 22.68 |
| sd | — | 7.52 | 2.76 | 6.86 | 3.30 | 1.10 | 3.04 |

## 4 Discussion

In this study, we used optically pumped magnetometers to record the magnetic field components of the propagating muscular action potential along the fibers of the abductor digiti minim muscle. In addition a bipolar surface and a monopolar fine wire emg and a bipolar fine wire emg was recorded. The muscle was activated by electric stimulation of the ulnar nerve with different stimulation strength. The stimulation strength was systematically altered to stimulate increasing amounts of neuro muscular units.

As expected the more neuro muscular units were stimulated the bigger the magnitude of the MMG signal and EMG signal became. The detailed statistical analysis showed a linear correlation between the magnitude of the MMG and EMG signal. With this finding it is plausible to apply the same knowledge that we have for the EMG signal to the MMG signal. As an example in this study it was shown how MMG could be used to predict the number of neuromuscular units.

Using computer simulations this finding could also be reproduced by the finite element simulation from (Klotz, 2022). In addition the simulation emphasizes the close correlation between the number of muscle fibers activated, the numbers of motor units and the magnetic signal strength.

The latest generation of OPM sensor allows even a recording of the magnetic field in all three spatial directions. This enabled us to reconstruct the precise spatial direction of the magnetic field vector generated by the muscle. Our data shows that the direction of the vectors is spatially stable and shows only a standard deviation of 5°. This finding highlights the possibilities of the MMG technique to reconstruct the location of the muscular action potential and may solve as fiducial marker for muscle vitality in the clinical setting.

While this study was quite successful, it had some critical limitations. The OPM devices used have a restricted bandwidth and therefore, fine, and fast components of the muscular action potential could not be recorded. It might possible that the fast components would show more variability with the number of motor units activated. Therefore, we plan to repeat this study with a sensor with a higher bandwidth. In addition, only one sensor was used. It might be more appropriate to use a sensor array located along the muscle in order to search for spatial differences in the longitudinal extend of the muscle.

## Data Availability

All data produced in the present study are available upon reasonable request to the authors.

## Abbreviations

MMG: magneto myography
OPM: optically pumped magnetometer
MAP: muscle action potential

## Footnotes

1 Bipolar EMG needles are ferromagnetic and therefore cannot be used with OPM devices.

---

**Supp. Figure 01.**
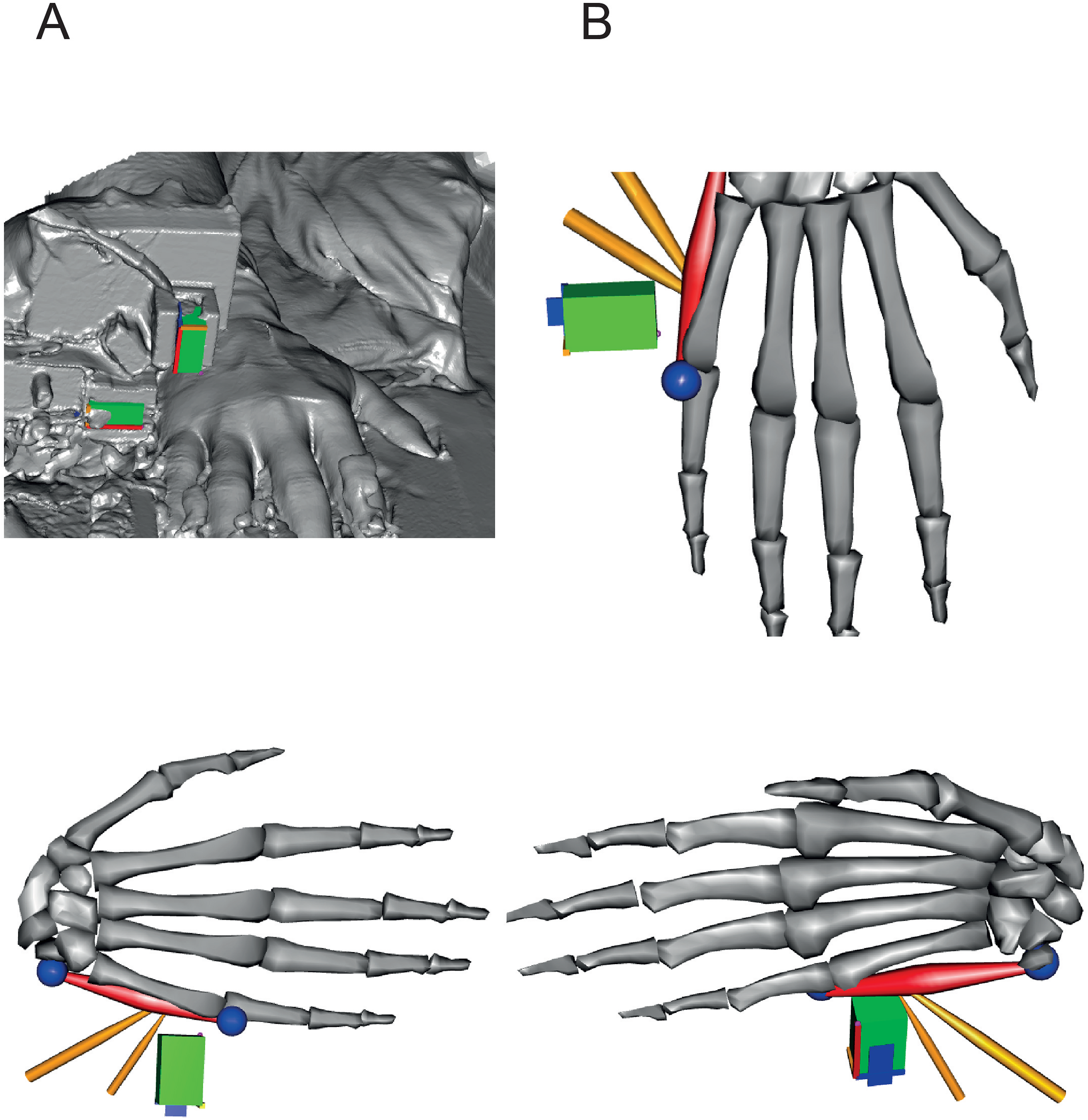

**Supplementary Figure 2 - Subj. 2.**
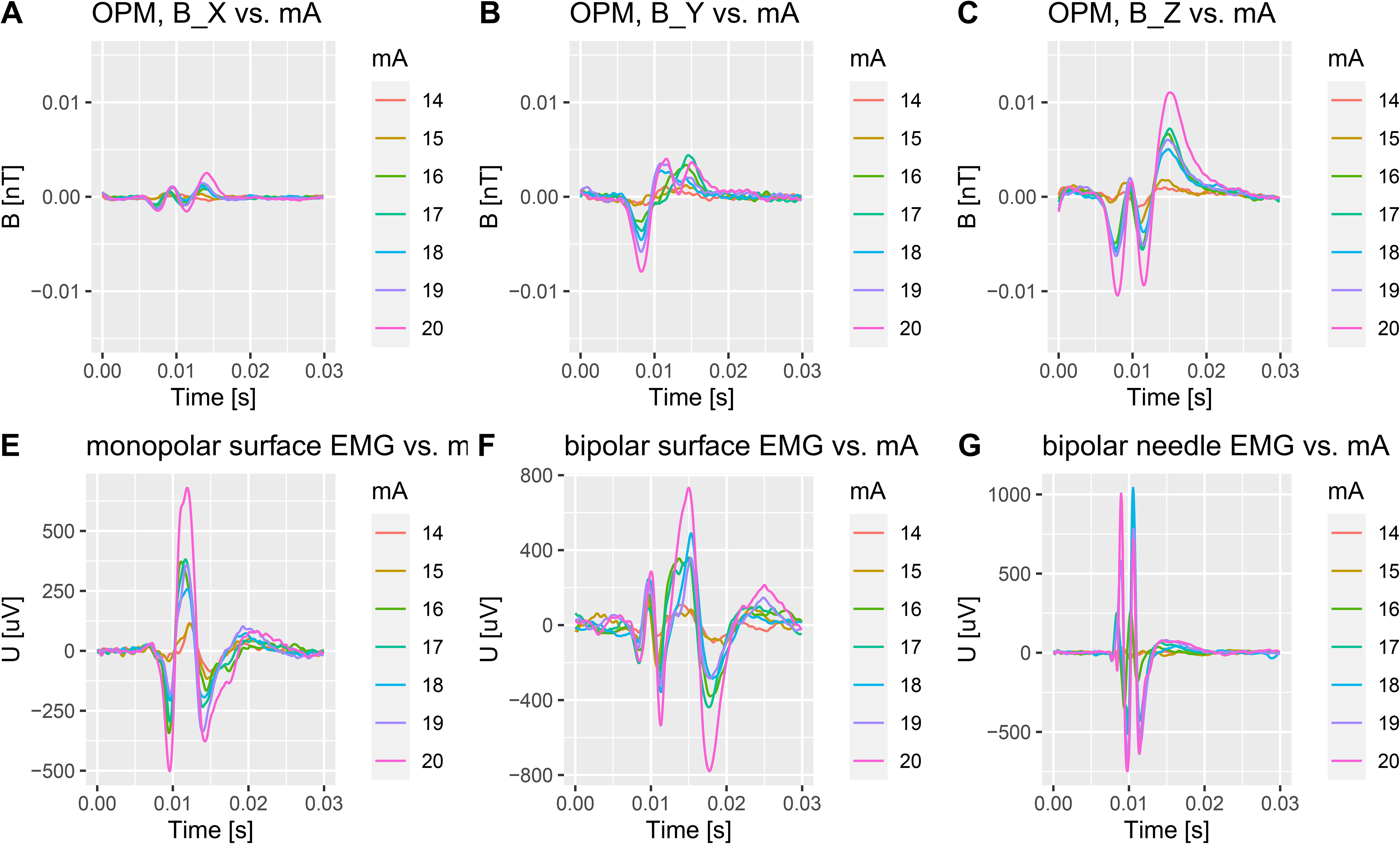

**Supplementary Figure 3.**
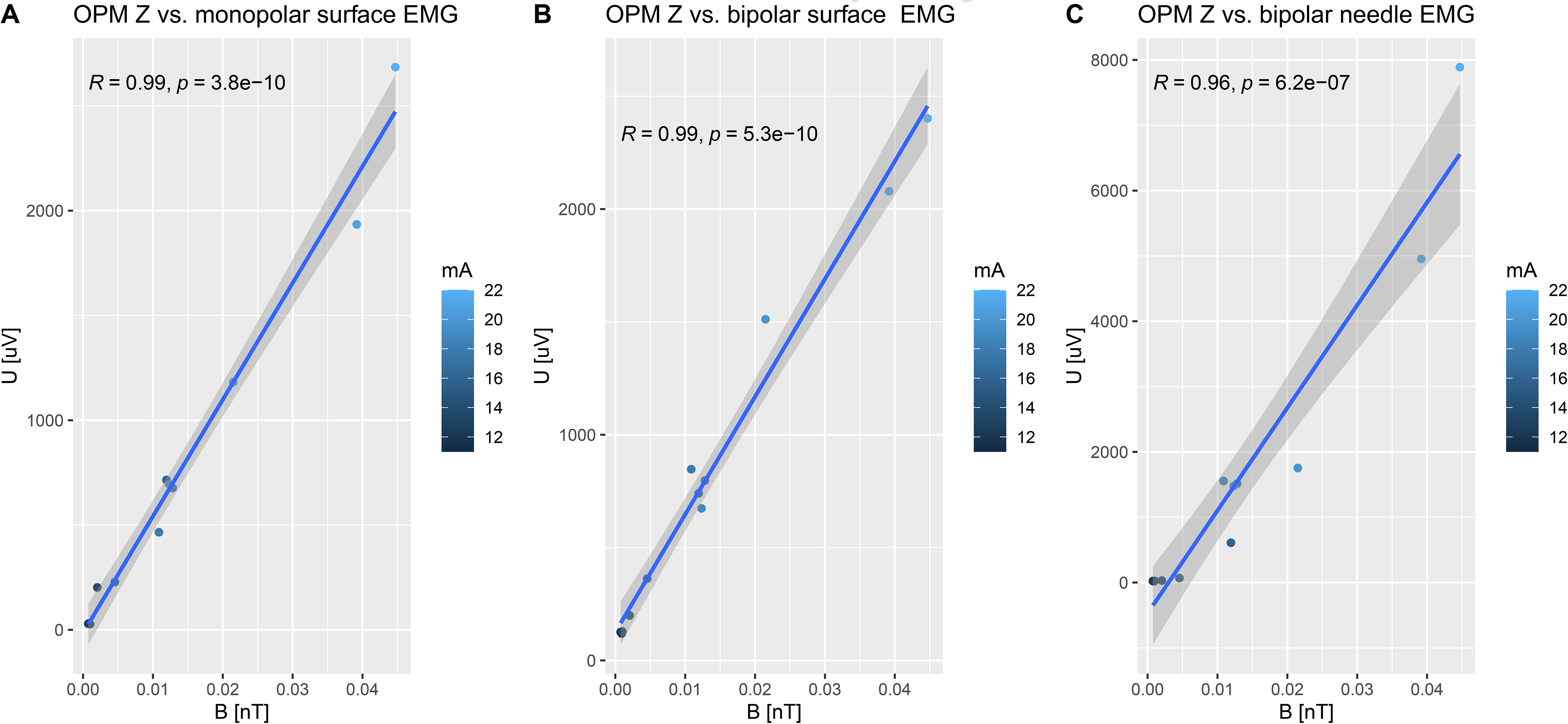

